# RetroTest unravels LINE-1 retrotransposition in Head and Neck Squamous Cell Carcinoma

**DOI:** 10.1101/2024.07.24.24310921

**Authors:** Jenifer Brea-Iglesias, Ana Oitabén, Sonia Zumalave, Bernardo Rodriguez-Martin, María Gallardo-Gómez, Martín Santamarina, Ana Pequeño-Valtierra, Laura Juaneda-Magdalena, Ramón García-Escudero, José Luis López-Cedrún, Máximo Fraga, José MC Tubio, Mónica Martínez-Fernández

**Author notes:** Corresponding author: Monica Martínez Fernández. Translational Oncology. Galicia Sur Health Research Institute (IIS Galicia Sur). Hospital Álvaro Cunqueiro. Estrada de Clara Campoamor, 341, 36213 Vigo, (Spain). Equal contribution (Jenifer Brea-Iglesias, Ana Oitabén). Present address: Centre for Genomic Regulation (CRG), The Barcelona Institute of Science and Technology, Barcelona (Spain). Universitat Pompeu Fabra (UPF), Barcelona (Spain).

## Abstract

The relevant role of LINE-1 (L1) retrotransposition in cancer has been recurrently demonstrated in recent years. However, their repetitive nature hampers their identification and detection, hence remaining inaccessible for clinical practice. Also, its clinical relevance for cancer patients is still limited. Here, we develop a new method to quantify L1 activation, called RetroTest, based on targeted sequencing and a sophisticated bioinformatic pipeline, allowing its application in tumor biopsies. First, we performed the benchmarking of the method and confirmed its high specificity and reliability. Then, we unravel the L1 activation in HNSCC according to a more extensive cohort including all the HNSCC tumor stages. Our results confirm that RetroTest is remarkably efficient for L1 detection in tumor biopsies, reaching a high sensitivity and specificity. In addition, L1 retrotransposition estimation reveals a surprisingly early activation in HNSCC progression, contrary to its classical association with advanced tumor stages. This early activation together with the genomic mutational profiling of normal adjacent tissues supports field cancerization process in this tumor. These results underline the importance of estimating L1 retrotransposition in clinical practice towards an earlier and more efficient diagnosis in HNSCC.

**Highlights:**

- RetroTest represents the first method to determine LINE-1 retrotransposition from tumor biopsies in real clinical settings.
- RetroTest not only offers global LINE-1 retrotransposition ratios but also identifies the active LINE-1 source elements.
- RetroTest elucidates a really early LINE-1 activation in early tumor stages of Head and Neck Squamous Cell Carcinoma.
- Whole Genome Analysis and LINE-1 retrotransposition demonstrates processes of field cancerization in Head and Neck Squamous Cell Carcinoma.
- LINE-1 retrotransposition favors an earlier and more efficient Head and Neck Squamous Cell Carcinoma diagnosis.

## 1. Introduction

Approximately half of human genome is composed of transposable elements (TEs), sequences with the ability of moving from one place to another, changing the normal structure of the genome in the places where they are integrated [1,2]. Among them, long interspersed nuclear element retrotransposons (LINE-1, L1) represent 17% of the entire DNA content with approximately 500,000 copies, most of them truncated or inactive [3–6]. Specifically, only a small subset of these L1s is active in the human genome, although they stay transcriptionally repressed due to epigenetic mechanisms that prevent the damage that their mobilization would cause [7]. When this repression is lost, L1 activation can cause different diseases, including cancer [8, 9].

In the framework of the International Consortium of PanCancer (PCAWG), our previous analyses shown that somatic L1 insertions represent the major restructuring source of cancer genomes, especially important for head-and-neck squamous cell carcinoma (HNSCC)[9]. Aberrant L1 retrotransposition contribute to instability within cancer genomes, promoting cancer-driving rearrangements that involve the loss of tumor-suppressor genes and/or the amplification of oncogenes, and favoring some cancer clones to survive and grow [9].

Despite this demonstrated impact of L1 activation in cancer genomes, the repetitive nature of L1 elements and their dispersion along the genome hinder their real activation estimates in tumor samples, preventing its translation into patient diagnosis/prognosis. This repetitive nature compels that the majority of the available detection methods are based on Whole Genome Sequencing (WGS), unfeasible for most hospitals in their diary practice, or require such good quality DNA input quantities that remain unaffordable for a clinical practice mainly based on small and degraded tissue biopsies.

HNSCC represents the second tumor type with highest L1 activation [9], but the clinical implications of L1 retrotransposition are still to be elucidated. Head and Neck Cancer is a heterogeneous group of cancers of which more than 90% are diagnosed as HNSCC, arising in the stratified epithelium of the oral cavity, pharynx, and larynx [10,11]. Tumor development is often triggered by chronic exposure to tobacco or alcohol, while infection with high-risk human papillomaviruses (HPVs) causes a substantial and rising proportion of these tumors [12]. The lack of symptoms in the early stages together with the non-existent cancer biomarkers lead towards most diagnoses at advanced stages, where the 5-year survival rate is less than 50% [11]. Thus, there is an urgent need to find molecular biomarkers that can facilitate an earlier diagnosis and increase patientś life expectancy.

Based on this, we aimed to unravel the impact of possible L1 activation along HNSCC development in a clinical setting. First, we have developed a new efficient method based on L1 transductions, RetroTest, to measure L1 activation even in biopsies with low DNA inputs. Then, we have evaluated RetroTest sensitivity and specificity, demonstrating its high values. Finally, we have assessed L1 retrotransposition along different tumor stages, unraveling for the fist time a really early activation in HNSCC and field characterization, arising L1 as a promising early diagnostic biomarker.

## 2. Materials and methods

### 2.1. Patients and tumor samples

Tumor samples and medical records were analyzed in a series of 96 HNSCC patients of 3 different cohorts (Complexo Hospitalario Universitario de A Coruña (CHUAC), Biobanco Vasco and Fundación Pública Galega de Medicina Xenómica (FPGMX)). The Ethical Committee for Clinical Research of Santiago-Lugo approved the study (CEIC 2018/567). Fresh-frozen and FFPE HNSCC tissue biopsies were collected, and characteristics of each tumor are specified in Table 1. The histopathologic status was confirmed by each corresponding Pathology Department following the latest TNM guidelines.

**Table 1.**
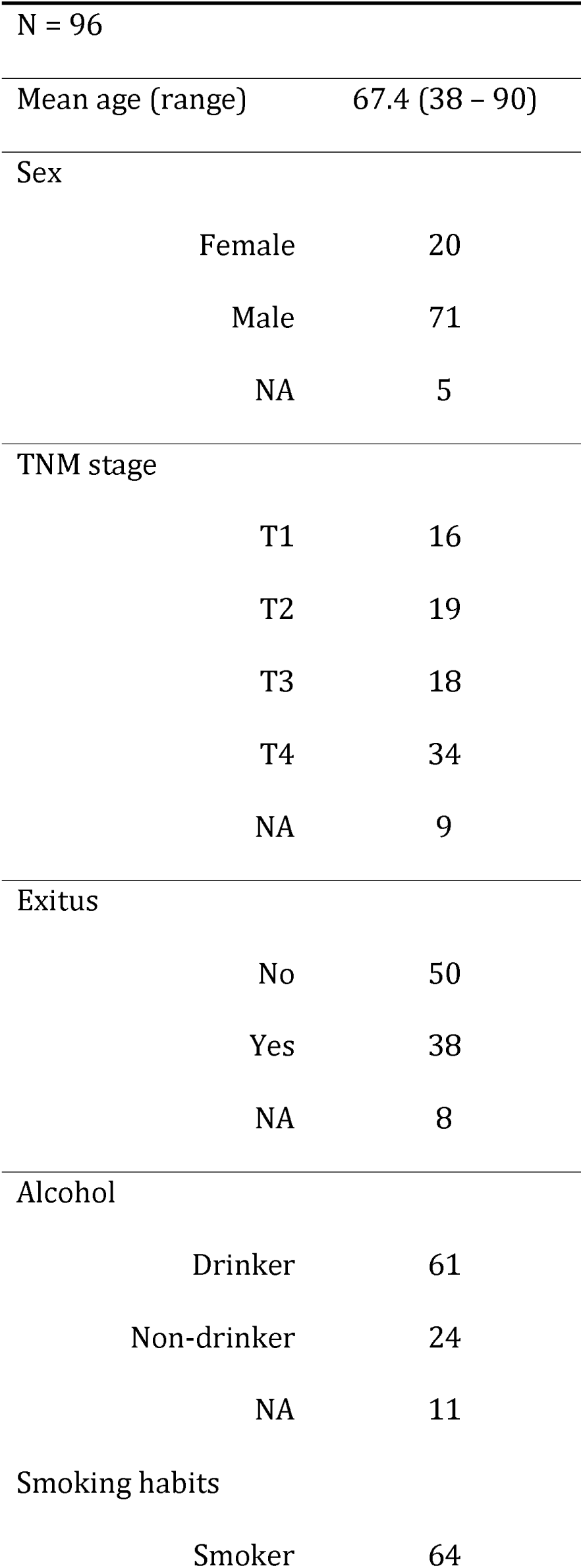
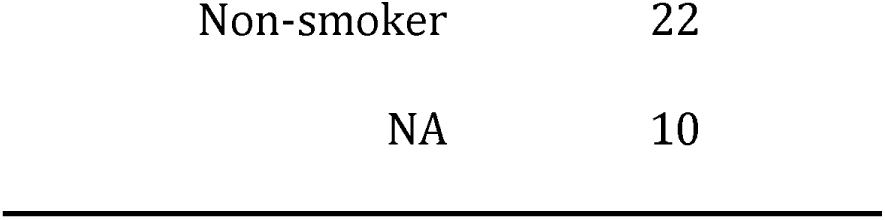
Baseline characteristics of the HNSCC patients and clinicopathological results in the series.

### 2.2. DNA isolation

Genomic DNA (gDNA) was extracted from fresh-frozen tissue and formalin-fixed paraffin-embedded (FFPE) samples using AllPrep DNA/RNA and AllPrep FFPE DNA/RNA Mini Kits (Qiagen), respectively. DNA quantification and integrity were assessed using Qubit dsDNA BR Assay Kit in Qubit 4.0 (ThermoFisher Scientific) and a 4200 TapeStation system (Agilent).

### 2.3. RetroTest method: design, library construction and target sequencing

During L1 transcription, the processing machinery sometimes bypasses the L1 polyadenylation signal until a second 3’ downstream polyadenylation site, mobilizing unique sequences downstream of the element in a process called L1 3’ transduction. This process has been reported to occur in around 10% of the L1 mobilizations [7] and can be used as an indirect measurement of real active L1 elements. Accordingly, there are three different types of retrotranspositions: solo-L1 (TD0), when a partial or complete L1 is retrotransposed; partnered transductions (TD1), in which a L1 and downstream unique sequence are retrotransposed; and orphan transductions (TD2), in which only the unique sequence downstream of the active L1 is mobilized without the associated L1 [7]. RetroTest is based on targeted sequencing, where the capture probes are designed against the unique sequence downstream of the 124 L1 full-length competent elements previously described [7] using SureSelect design from Agilent. RetroTest identifies L1 TD2 orphan insertions through the detection of discordant reads and clipped reads. The main idea behind the method is that discordant read pairs, where one of the mates is mapped to a L1 3’ downstream sequence while the other is mapped to the insertion target sequence, support an insertion. In addition, clipped reads, mapped to the target sequence but containing a discordant extreme blatting to a L1 3’ downstream sequence, are detected to identify the breakpoint of the insertion (Fig. 1A). Here, a minimum size of 2 reads per cluster and a minimum of 4 supporting reads was specified to call a L1 insertion (Additional file 1). Target libraries were constructed with SureSelect Target Enrichment System for Illumina Paired-End Multiplexed Sequencing (Agilent). 100 ng of gDNA from each sample were sheared using a Covaris M220 Focused-Ultrasonicator (Covaris) and libraries and capture with targeted RNA baits were performed. The multiplexed samples were sequenced with Illumina 150bp paired-end.

**Figure 1.**
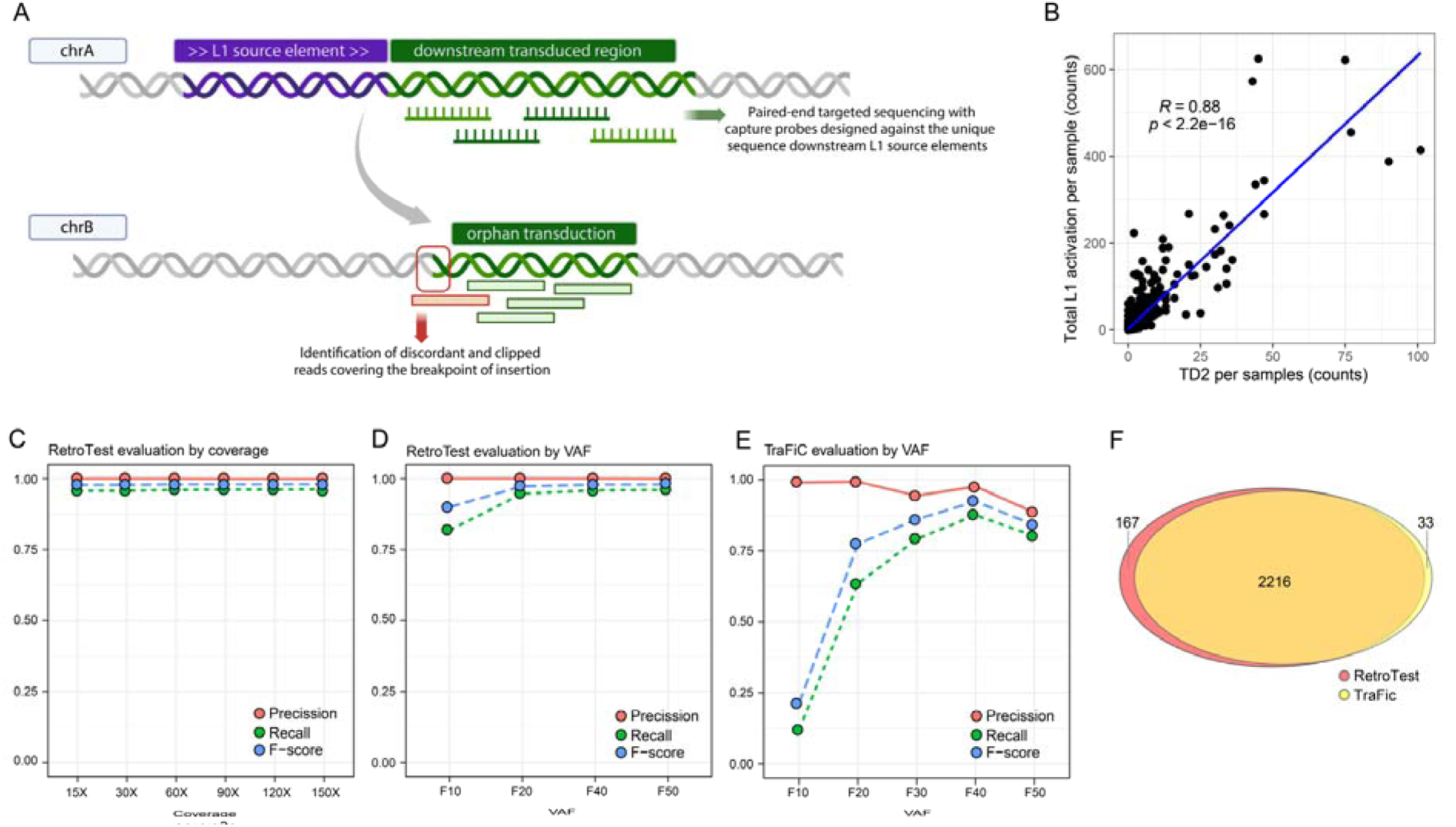
RetroTest design and benchmarking. A. RetroTest design. B. Scatterplot and correlation between L1 activation and orphan transductions (TD2) in the International Consortium of PanCancer (PCAWG) data. The number of L1 3’ transductions measured L1 activation. C. Performance of RetroTest for different sequencing coverages using an artificially generated GRC37/hg19 genome with a total of 2,480 randomly distributed L1 transductions at 50% VAF. D. Performance of RetroTest with respect to the VAF of L1 integrations, using the artificially generated GRC37/hg19 genome with a total of 2,480 randomly distributed L1 transductions at different VAFs. E. Performance of TraFiC with respect of the VAF of L1 integrations, using the artificially generated GRC37/hg19 genome with a total of 2,480 randomly distributed L1 transductions at different VAFs. F. Venn diagram of the number of L1 insertions detected by RetroTest and TraFiC in the artificial genome with a VAF of 50% for L1 insertions.

Sequencing reads were mapped to the hg19 reference genome by Burrows-Wheeler Aligner BWA-mem [13]. Samtools [14] was used to sort the aligned reads and to index the obtained bam file, applying Bammarkduplicates2 from Picard tools [15] to mark duplicated reads.

### 2.4. RetroTest benchmarking

We generated simulated paired-end read datasets using ART [16] and several commands from MEIGA-MEIsimulator, an in-house bioinformatic tool (Additional file 1).

To mimic the capture process, we selected read pairs with at least one of the mates mapping on a given set of target regions with Picard FilterSamReads v2.18.14 [15]. We used the resulting bam files to assess sensitivity and specificity under different conditions. To test how our method performs with subclonal events, we simulated transductions at different VAFs (10%, 20%, 40% and 50%). We also studied how coverage affects the detection power of our algorithm. Using Picard DownSampleSam v2.18.14 [15], we subsampled reads from a 150x simulation at 50% VAF under different sequencing depths (15x, 30x, 60x, 90x, 120x and 150x). We compared RetroTest performance with Transposon Finder in Cancer (TraFiC), used previously to explore somatic retrotransposition in PCAWG [7]. Both were used to call the simulated events. Precision was calculated dividing the number of true positive calls by the number of total calls performed by the method. Recall was calculated dividing the number of true positive calls by the number of total simulated events. True positives, False positives and False negatives were identified by intersecting the coordinates of simulated events with the coordinates of the calls using BEDTools intersect [17]. To compare RetroTest and TraFiC results, Venn diagrams with the insertions detected by each method were plotted by using vennDiagram R package.

### 2.5. Whole Genome Sequencing and determination of mutation profile

To obtain WGS data, DNA was sent to an external service (Macrogen). Truseq Nano DNA Libraries (350bp) were constructed and sequenced in a NovaSeq6000 Illumina platform (150bp paired-end). For a detailed explanation see Additional File 1.

### 2.6. Statistical analyses

The association between L1 transduction rate (corrected by coverage) and patient clinical features was assessed by multiple linear regressions. Wilcoxon or Fisher test, depending on sample size, were applied to compare the differences in mean values for clinical variables. Overall survival (OS), progression free survival (PFS) and survival probability analyses were performed with the survminer and survival R packages, using log-rank test to compare different groups. The association between survival and clinical variables was evaluated by Cox regression.

### 2.7. Enrichment analysis

All enrichment analyses were performed using enrichR R package [18]. The complete list of pathways databases can be found in Additional File 1.

## 3. Results

### 3.1. RetroTest Benchmarking

During L1 transcription, transductions are reported to occur in around 10% of the L1 mobilizations [7], moving also unique sequences downstream of the source element. RetroTest is designed to capture these mobilized and downstream-transduced unique sequences from orphan transductions (TD2) and use them as barcodes (Fig. 1A). These barcodes can identify unequivocally the insertions caused by these 124 L1 source elements active in cancer [7,9]. With this aim, we focused RetroTest probe design on the first 5000 nucleotides adjacent to the L1 3’ regions for each of the 124 L1 source elements, since these are the regions most frequently transduced [7]. Since RetroTest uses transductions as an indirect measure of real L1 activation, we first compared L1 activation versus TD2 using the whole PCAWG data. In this way, we confirmed a high and statistically significant correlation between both events (r=0.88, p=2.2e^-16^), and a relation of 1:10 as previously described (Fig. 1B).

Next, we optimized the lab protocol for both FFPE and fresh-frozen tissues and developed an associated bioinformatic pipeline. We evaluated the performance and accuracy of RetroTest in detecting L1 activation by generating an artificial cancer genome. In this genome, we have randomly distributed a total of 2480 L1 transductions. Using simulations, we assessed the performance of RetroTest for different sequencing depth: 15x, 30x, 60x, 90x, 120x and 150x. RetroTest obtained a precision around 0.99 in all cases and a recall around 0.96 (Fig. 1C). We also evaluated the variation of the performance depending on the variant allele frequency (VAF) of the integrations, precisely for the following VAFs: 10%, 20%, 40% and 50%. In this case, RetroTest obtained a precision around 1, decreasing for lower VAFs, and a recall ranged from 0.81 to 0.96, augmenting as increasing the VAF (Fig. 1D).

Then, we compared RetroTest and the classical TraFiC, used for WGS in PCAWG [7]. As TraFiC is designed to work only with standard WGS 30x data, we compared the analysis varying exclusively the VAFs. TraFiC precision ranged from 0.99 to 0.88 and recall ranged from 0.11 to 0.87 as VAF increased (although the maximum recall was obtained for VAFs of 40%, being 0.87) (Fig. 1E) (Additional File 2: Table S1). We compared the performance of both methods by intersecting both calls in the mock tumor genome. Using a VAF of 50%, most of the variants were called by both methods, concretely 2216 variants, while RetroTest exclusively called 167, and TraFiC 33 private events, most of them resulting in false positives according to IGV (Fig. 1F).

### 3.2. L1 activation in HNSCC

Once confirmed the accuracy and precision of RetroTest, we decided to apply it to a more extensive HNSCC cohort, composed of 96 tumors along the tumor stages, from T1 to T4 (Table 1). We detected L1 activation in 75% of the patients (Fig. 2A), out of which 48.6% showed high activity (beyond the median) (Table 2). When the activation was studied along the different tumor stages of the disease, advanced disease (T3-T4) showed statistically higher L1 activation compared with early stages (T1-T2) (p=0.0072) (Fig. 2B). Following tumor staging, L1 activation was detected in 62.5% of T1 tumors, 63.1% of T2 tumors, 94.4% of T3 tumors, and 73.5% of the tumors in T4 (Table 2). The detection of L1 activation in all the tumor stages among the different patients, even in the first stage of the disease (T1), supports an early L1 activation during tumor progression.

**Figure 2.**
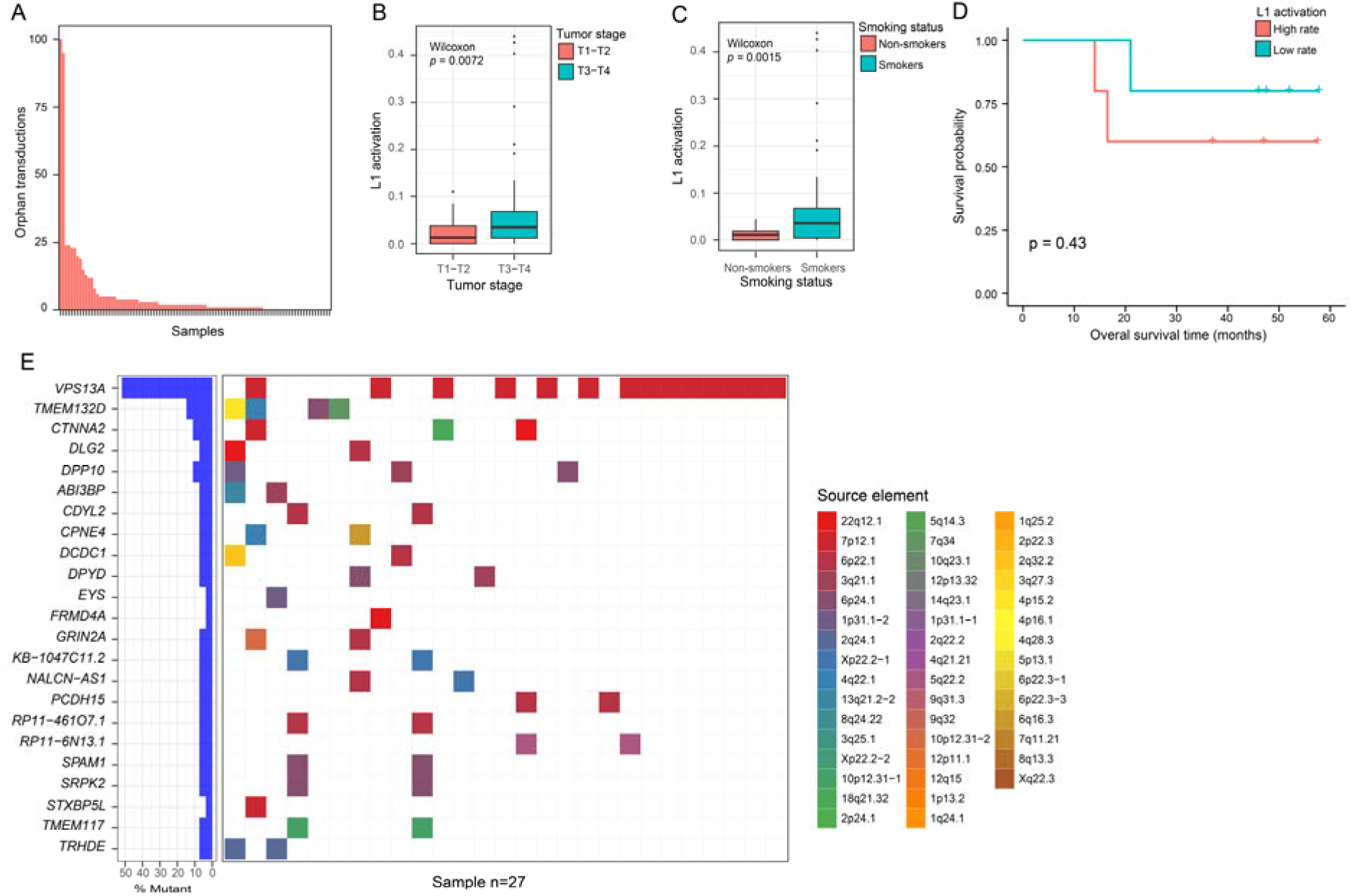
L1 activation measured by RetroTest in the HNSCC cohort (n=96). A. Quantification of L1 activation in HNSCC tumors as the number of orphan transduction detected by RetroTest. B. Boxplot of L1 activation with respect to early (T1-T2) and advanced (T3-T4) TNM stages. Differential activation p-value was derived by the Wilcoxon test. To correct coverage-related bias, L1 activation was calculated as the number of TD2 divided by its median coverage. C. Boxplot of L1 activation with respect to smoking status. Differential activation p-value was derived by the Wilcoxon test. To correct coverage-related bias, L1 activation was calculated as the number of TD2 divided by its median coverage. D. Kaplan-Meier curves for overall survival with respect to L1 activation rate at T1 stage. Patients were grouped into high (above the median) and low (below the median) L1 activation rate. Log-rank test was used to calculate the p-value. To correct coverage-related bias, L1 activation was calculated as the number of TD2 divided by its median coverage. E. Oncoplot showing the genes affected by L1 insertions and their original source elements. A total of 27 patients presented genes affected by L1 insertions from 46 different source elements.

**Table 2.**
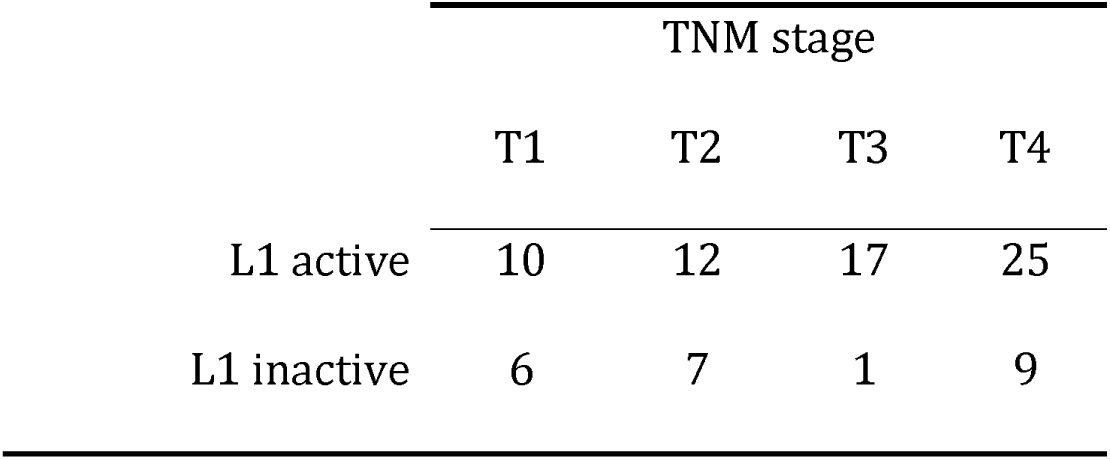
Number of HNSCC patients showing L1 activation along TNM stages.

|  | TNM stage |  |  |  |
| --- | --- | --- | --- | --- |
|  | T1 | T2 | T3 | T4 |
| L1 active | 10 | 12 | 17 | 25 |
| L1 inactive | 6 | 7 | 1 | 9 |

Then, we analyzed L1 activation considering different patients’ clinical characteristics, finding no statistically significant association between the L1 activation and alcohol consumption (p=0.14) or sex (p=0.055). Interestingly, smoker patients showed statistically significantly higher L1 activity than non-smokers (p=0.0015) (Fig. 2C). We did not find an association between L1 activation and survival probability (p=0.59 active vs. inactive, p= 0.49 high vs. low rate) (Additional File 3: Fig. S1), although when considering those T1 patients who have L1 already active at early stages, Kaplan-Meier curve showed that they tend to have a lower survival probability, not being statistically significant (p=0.43) (Fig. 2D).

Finally, in addition to localizing the position of each transduction, RetroTest can also identify the source L1 element that has been mobilized. Thus, we have detected L1 transductions in 23 genes throughout 27 patients. As shown in Fig. 2E, most of these insertions are caused by a few source elements that resulted very active in HNSCC, especially the 22q12.1.

### 3.3. HNSCC mutation profile and L1 activation

To further characterize the molecular profile of L1 activation in our HNSCC cohort, we obtained WGS data from 19 tumor samples from patients measured by RetroTest. To detect only somatic variation, their corresponding paired-normal samples were included as germline control. We detected a median of 40 single nucleotide variants (SNVs) and INDELs, identifying a total of 1012 somatic variants throughout the cohort, affecting 918 genes (Additional File 2: Table S2). As shown in Fig. 3A, we did not detect a correlation between L1 activation and the general tumor mutation burden (TMB). Our results showed that the most frequently mutated gene was TP53 (36.8%), followed by NOTCH1 (26.3%), MT-ND5 (26.3%), FAT1, and GRIN2A (21.1%). Interestingly, we found that most of the patients with TP53 mutations showed also high L1 activity (71.4%). In fact, when we compared the L1 activation with the TP53 mutation, we found a clear association (p=0.056) (Fig. 3B). Enrichment analyses with the mutated genes showed involved key processes for cancer progression such as Notch signaling, TGFß signaling, and again p53 activity regulation (Fig. 3C) (Additional File 2: Table S3). CHEA analysis revealed the alteration of transcription factors related to epigenetic mechanisms including Polycomb members (EZH2, SUZ12) (Fig. 3D) (Additional File 2: Table S4).

**Figure 3.**
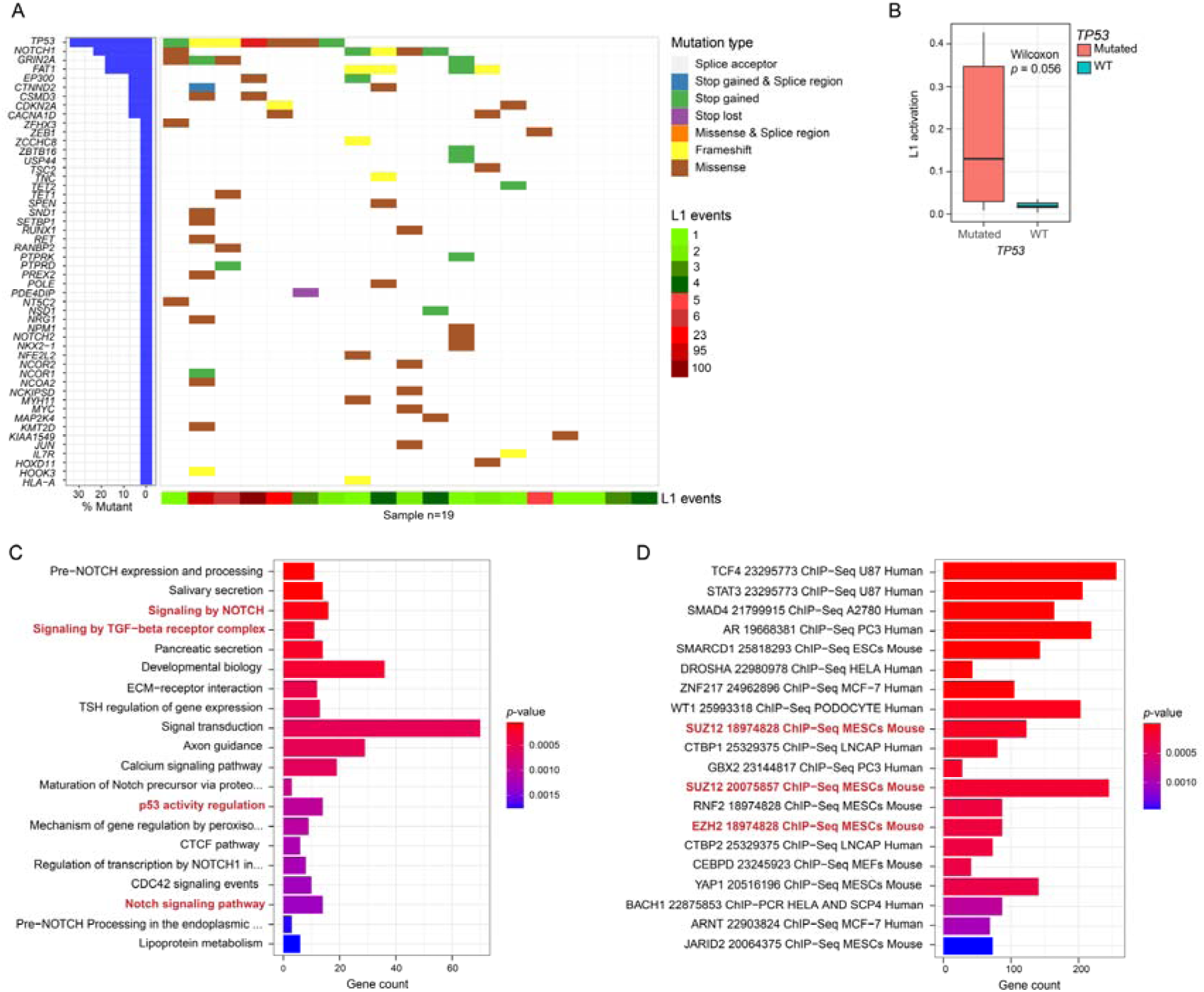
Characterization of the mutational profile of HNSCC patients by WGS (n=19). A. Oncoplot showing the genes harboring somatic mutations in HNSCC patients. The type of mutation in each gene and the L1 activation (number of L1 insertions in each patient) is shown. B. Boxplot of L1 activation with respect to TP53 mutation or wild-type status. To correct coverage-related bias, L1 activation was calculated as the number of TD2 divided by its median coverage. Differential activation p-value was derived by the Wilcoxon test. C. Barplot of the Pathway enrichment analysis based on the 918 genes somatically mutated genes. Enrichment p-values were calculated with the Fisher exact test. D. Barplot of the transcription factor binding enrichment analysis based on the 918 genes somatically mutated genes. Enrichment p-values were calculated with the Fisher exact test.

### 3.4. Profiling normal adjacent tissues to the HNSCC tumor (NAT)

To further understand the early HNSCC features, we decided to evaluate a possible field cancerization process, in which the normal cell population is replaced by cancer-primed cells, without anatomical or morphological changes, but being already premalignant at molecular level. To this, we analyzed the available normal samples obtained from adjacent tissues to the tumor and the peripheral blood mononuclear cells (PBMCs), used as germline control (Fig. 4A). We detected a total of 25 high impact and/or possibly pathogenic somatic variants affecting the NAT (Additional File 2: Table S5). Most of these specific mutations (n=20; 80%) were exclusive from NAT tissue, including those affecting key genes such as NOTCH1 (the most mutated gene), FAT1 or PPARD; while 20% were shared between NAT and tumor tissue, affecting genes such as CDKN2A.

**Figure 4.**
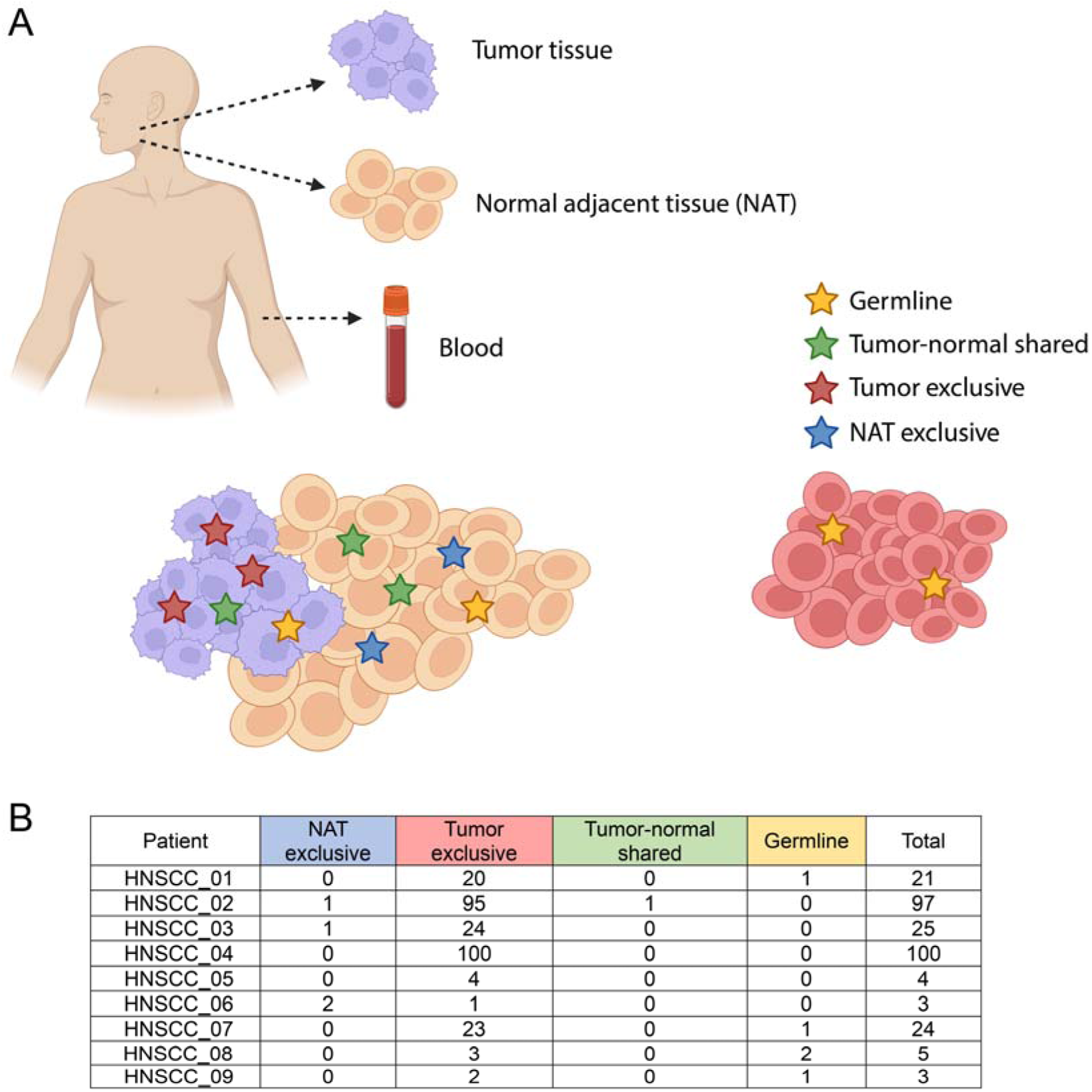
Evaluation of L1 activation in normal adjacent tissue. A. Schematic representation of the evaluation of the field cancerization process. B. Number of L1 active elements in normal adjacent tissue of HNSCC patients (n=9), compared to L1 activation in paired tumor tissue and PBMCs as germline control from the same patients.

To elucidate whether L1 further supports field cancerization, we evaluated the NAT of a total of 9 patients by RetroTest. We compared the L1 elements active in the tumor, in the NAT, and in their corresponding paired germline. In this way, we could confirm that most of the L1 activation was present only in the tumor, 5 insertions resulted germline, and 1 element appeared active and shared by tumor and NAT. Surprisingly, 4 insertions appeared exclusively in NAT (Fig. 4B). Thus, we could confirm field cancerization and demonstrate that L1 is already active in NAT, supporting once again its early activation in HNSCC.

## 4. Discussion

Since the recently demonstrated high impact of L1 in cancer genomes [7,9], L1 has been evaluated as a cancer biomarker in different studies assessing its activity by evaluating L1 methylation, RNA expression, or protein levels [19–22]. However, most of L1 genome sequences are truncated and not functional, so results can present important biases in their L1 estimations, while its translation into clinical routine can be challenging due to RNA/protein instability. Previous proposed technologies based on DNA full-length L1 retrotransposon capture sequencing, such as RC-seq [23], present the associated possible bias of the real L1 activation, besides only offer global L1 estimates, and important DNA requirements (2.5μg starting genomic DNA), unaffordable for clinical practice. The most recent approaches are based on the identification of L1 insertions as real L1 activation measure (TraFiC [7], xTea [24], MELT [25] and Mobster [26]). They use Whole Genome or Whole Exome Sequencing from Illumina pair-end short reads, nevertheless, short reads hamper the detection of transposable elements insertions in highly repetitive or complex rearrangement regions. Additionally, these high throughput approaches are not affordable for most of the hospitals. More recently, long reads sequencing has arisen as a new possibility (xTea [24], PALMER [27]), but again their requirements of huge amounts of high-quality DNA remain unaffordable for biopsies mainly composed of small samples and fragmented DNA. Thus, incorporating L1 activation into the clinical practice requires new methods supported by standardization and rigorous validation to demonstrate its utility.

Here, we present RetroTest: a new method to detect real L1 activation in clinical samples with low DNA input requirements, both from fresh/frozen or FFPE biopsies. Its novelty and power lie in the targeted detection of active source L1 elements, which showed a highly and direct statistical correlation with total L1 activity (as demonstrated by our data) in a more cost-effective than previously proposed approaches. Our method not only offers global L1 estimates but also identifies the L1 elements active in each real sample. Our benchmarking supports the high precision (1) and recall of RetroTest (0.81-0.96), improving according to coverage, even reaching the possibility of detecting subclonal insertions. The high coincidence in the variants called by TraFic and RetroTest, in both simulations and real-world data, supports the high potential for this new methodology.

L1 somatic retrotransposition was previously demonstrated as the second most frequent type of structural variants among HNSCC genomes [9]. Accordingly, our results demonstrate that most HNSCC patients (75%) present L1 activation, even near half of them with high levels (49%). The activation was higher in late stages, as found in Barret’s esophagus, where lower L1 activity was detected in the early stages increasing with cancer progression [28]. Interestingly, we found a surprisingly early activation already in the first stages of HNSCC, with activation of L1 in 62-63% of the T1 patients. These data pointed towards L1 activation as an early event in the configuration of HNSCC genomes and, thus, in the development of the disease. In fact, those stage T1 patients with L1 activation tended to present a lower OS.

We found an association between high L1 activity and smoking habits. Previous studies had reported higher hypomethylation rates of L1 in smokers [29], even in non-cancerous epithelial tissues [30]. Considering that around 75% of HNSCC are associated with tobacco [31], this mechanism could be responsible for the high L1 activity levels in HNSCC, since methylation is one of the best-demonstrated mechanisms preventing L1 reactivation [7,32]. In fact, L1 hypomethylation has been associated with worse prognosis in HNSCC, including a higher risk of relapse [33–35]. This hypomethylation has been found as an early event in CRC, gastric and oral cancer [36–39]. Interestingly, it was reported that L1 methylation levels were significantly lower in oral premalignant lesions of patients who then developed oral cancer [39].

The source element L1 most active in our cohort was that at 22q12.1, coincident with our previous results [7, 10]. This L1 is located antisense to an intron of the TTC28 gene and has been also recently identified as the intact LINE-1 mRNA most highly expressed in breast, ovarian and colon cancer [40], and the L1 element accounting for most transductions in colorrectal cancer [33], supporting the same hottest activity in HNSCC.

Our WGS analyses confirmed no correlation between L1 activity and TMB, but an association between TP53 mutation and L1 activation, in line with previous studies suggesting that TP53 can repress L1 mobilization [9,40–43]. We also identified an important epigenetic regulation among the transcription factors strongly interacting with the mutated genes, specially related to repressive complex Polycomb. Intriguingly, Mangoni et al. have just deciphered that L1 RNAs can act as long non-coding RNAs and directly interact with the Polycomb during brain development and evolution [44]. We also reported previously that Polycomb could regulate lncRNA HOTAIR in bladder cancer [45]. In the same sense, Ishak et al. demonstrated an EZH2-dependent silencing of genomic repeat sequences, including L1 elements [46]. Therefore, additional analyses would be required to further address if this epigenetic network plays a role in cancer genome reorganization.

Then, to evaluate possible HNSCC early diagnosis potential biomarkers is fundamental to get insights into the transition from pre-tumoral to cancer disease. Thus, we evaluated the presence of field cancerization and identified genetic features that can eventually lead to cancer development [47], finding exclusive somatic mutations in the NAT and a small proportion shared with the tumor. Several studies have recently described patchworks of different clones in normal tissues, some of them even bearing driver mutations, increasing with age or smoking habits [48–50]. We found NOTCH1 and FAT1 as the most mutated genes, as Martincorenás results in normal skin and esophagus [48,51]. These genes showed also high mutation rates in the tumor, indicating the presence of a precancerous or cancer invasion field.

Finally, when we evaluated L1 activity, several mobilizations were exclusively found in NAT, and only one appeared shared with the tumor, again supporting a cancerization field. Somatic L1 retrotransposition events have been recently described in normal urothelium and colorectal epithelium, although with much lower rates than in bladder and colorectal cancer [52,53]. L1 activity has been also described in a few pre-tumor samples including Barrett’s esophagus and colorectal adenomas [8,28,54]. Therefore, L1 activation arises as a reinforced early event, even in pre-tumor stages, in the natural history of HNSCC, with the associated potential of L1 as a field cancerization biomarker.

## Conclusions

We present the development and benchmarking of RetroTest, which allows an easy measurement of real L1 activation from small tumor biopsies with high efficacy, favoring its implementation in real clinical settings. RetroTest revealed that most of the HNSCC patients present L1 activation, associated with smoking habits and already active in early stages of the disease and NAT, supporting a field cancerization process and its potential as early diagnostic biomarker.

## Data Availability

Whole Genome Sequencing data from the HNSCC cohort is deposited into the Sequence Read Archive (SRA) repository under the following BioProject ID: PRJNA1053897. RetroTest pipeline is publicly available at https://gitlab.com/mobilegenomesgroup/RETROTEST.

## Acknowledgements

Authors thank all the enrolled patients and their families. This research project was made possible through the access granted by the Galician Supercomputing Center (CESGA) to its supercomputing infrastructure. The supercomputer FinisTerrae III and its permanent data storage system have been funded by the Spanish Ministry of Science and Innovation, the Galician Government and the European Regional Development Fund (ERDF).

This work was supported by the Instituto de Salud Carlos III (ISCIII) and the European Social Fund (“Investing in your future”) (PI19/01113, and partially by P121/00208, co-funded by FEDER and the European Union), and the Spanish Association Against Cancer Scientific Foundation (IDEAS19122MART). A.O. was supported by a predoctoral fellowship from the Galician Innovation Agency, Xunta de Galicia (ED481A-2020/214). M.M.-F. and J.B. were previously supported by the Spanish Association Against Cancer Scientific Foundation (INVES207MART and PRDCR19007BREA_001, respectively). M.M.-F. is currently supported by the Miguel Servet program (CP20/00188) from the Instituto de Salud Carlos III (ISCIII) and the European Social Fund (“Investing in your future”). M.G-G. is supported by a postdoctoral fellowship from the Galician Innovation Agency, Xunta de Galicia (IN606B-2024/014).

## Ethics statement

The Ethical Committee for Clinical Research of Santiago-Lugo gave ethical approval for this work (CEIC 2018/567).

## Consent for publication

All authors give consent for the publication of this manuscript.

## CRediT authorship contribution statement

**Jenifer Brea-Iglesias and Ana Oitabén**: Conceptualization, Methodology, Software, Data curation, Investigation, Validation, Visualization, Writing – review and editing. **Sonia Zumalave**: Conceptualization, Investigation, Methodology, Software, Validation, Writing – review and editing. **Bernardo Rodríguez-Martin**: Conceptualization, Methodology, Software. **María Gallardo-Gómez**: Conceptualization, Visualization, Writing – review and editing. **Martin Santamarina**: Methodology, Validation. **Ana Pequeño-Valtierra**: Conceptualization, Investigation. **Laura Juaneda-Magdalena**: Methodology, Conceptualization, Writing-review and editing. **Ramón García-Escudero**: Methodology, Writing-review and editing. **Jose Luis López-Cedrún**: Methodology. **Máximo Fraga**: Funding acquisition. **José MC Tubio**: Conceptualization, Funding acquisition. **Mónica Martínez-Fernández**: Writing – review & editing, Writing – original draft, Supervision, Resources, Investigation, Project administration, Funding acquisition, Conceptualization. All authors approved the final version.

## ADDITIONAL FILES

**Additional file 1 (pdf): Supplementary Methods:** RetroTest library and target sequencing; RetroTest method in detail; RetroTest benchmarking; Whole Genome Sequencing and determination of mutation profile; Gene pathway databases uses for enrichment analysis

**Additional file 2 (xls): Table S1:** RetroTest benchmarking. **Table S2**: HNSCC somatic variants. **Table S3**: Gene pathways affected by HNSCC somatic variants. **Table S4**: Transcription factors binding to genes affected by HNSCC somatic variants. **Table S5**: HNSCC NAT somatic variants

**Additional file 3 (pdf). Figure S1**. Kaplan-Meier curves for overall survival in HNSCC cohort with respect to (A) L1 activation status (active vs inactive) and (B) L1 activation rate (with respect to the median, being high above vs low as below the median). Log-rank test was used to calculate the p-value.

## References

[1] H.H. Kazazian, J. V. Moran, The impact of L1 retrotransposons on the human genome, Nat. Genet. 19 (1998) 19–24. 10.1038/NG0598-19.

[2] E.S. Lander, L.M. Linton, B. Birren, C. Nusbaum, M.C. Zody, J. Baldwin, K. Devon, K. Dewar, M. Doyle, W. Fitzhugh, R. Funke, D. Gage, K. Harris, A. Heaford, J. Howland, L. Kann, J. Lehoczky, R. Levine, P. McEwan, K. McKernan, J. Meldrim, J.P. Mesirov, C. Miranda, W. Morris, J. Naylor, C. Raymond, M. Rosetti, R. Santos, A. Sheridan, C. Sougnez, N. Stange-Thomann, N. Stojanovic, A. Subramanian, D. Wyman, J. Rogers, J. Sulston, R. Ainscough, S. Beck, D. Bentley, J. Burton, C. Clee, N. Carter, A. Coulson, R. Deadman, P. Deloukas, A. Dunham, I. Dunham, R. Durbin, L. French, D. Grafham, S. Gregory, T. Hubbard, S. Humphray, A. Hunt, M. Jones, C. Lloyd, A. McMurray, L. Matthews, S. Mercer, S. Milne, J.C. Mullikin, A. Mungall, R. Plumb, M. Ross, R. Shownkeen, S. Sims, R.H. Waterston, R.K. Wilson, L.W. Hillier, J.D. McPherson, M.A. Marra, E.R. Mardis, L.A. Fulton, A.T. Chinwalla, K.H. Pepin, W.R. Gish, S.L. Chissoe, M.C. Wendl, K.D. Delehaunty, T.L. Miner, A. Delehaunty, J.B. Kramer, L.L. Cook, R.S. Fulton, D.L. Johnson, P.J. Minx, S.W. Clifton, T. Hawkins, E. Branscomb, P. Predki, P. Richardson, S. Wenning, T. Slezak, N. Doggett, J.F. Cheng, A. Olsen, S. Lucas, C. Elkin, E. Uberbacher, M. Frazier, R.A. Gibbs, D.M. Muzny, S.E. Scherer, J.B. Bouck, E.J. Sodergren, K.C. Worley, C.M. Rives, J.H. Gorrell, M.L. Metzker, S.L. Naylor, R.S. Kucherlapati, D.L. Nelson, G.M. Weinstock, Y. Sakaki, A. Fujiyama, M. Hattori, T. Yada, A. Toyoda, T. Itoh, C. Kawagoe, H. Watanabe, Y. Totoki, T. Taylor, J. Weissenbach, R. Heilig, W. Saurin, F. Artiguenave, P. Brottier, T. Bruls, E. Pelletier, C. Robert, P. Wincker, A. Rosenthal, M. Platzer, G. Nyakatura, S. Taudien, A. Rump, D.R. Smith, L. Doucette-Stamm, M. Rubenfield, K. Weinstock, M.L. Hong, J. Dubois, H. Yang, J. Yu, J. Wang, G. Huang, J. Gu, L. Hood, L. Rowen, A. Madan, S. Qin, R.W. Davis, N.A. Federspiel, A.P. Abola, M.J. Proctor, B.A. Roe, F. Chen, H. Pan, J. Ramser, H. Lehrach, R. Reinhardt, W.R. McCombie, M. De La Bastide, N. Dedhia, H. Blöcker, K. Hornischer, G. Nordsiek, R. Agarwala, L. Aravind, J.A. Bailey, A. Bateman, S. Batzoglou, E. Birney, P. Bork, D.G. Brown, C.B. Burge, L. Cerutti, H.C. Chen, D. Church, M. Clamp, R.R. Copley, T. Doerks, S.R. Eddy, E.E. Eichler, T.S. Furey, J. Galagan, J.G.R. Gilbert, C. Harmon, Y. Hayashizaki, D. Haussler, H. Hermjakob, K. Hokamp, W. Jang, L.S. Johnson, T.A. Jones, S. Kasif, A. Kaspryzk, S. Kennedy, W.J. Kent, P. Kitts, E. V. Koonin, I. Korf, D. Kulp, D. Lancet, T.M. Lowe, A. McLysaght, T. Mikkelsen, J. V. Moran, N. Mulder, V.J. Pollara, C.P. Ponting, G. Schuler, J. Schultz, G. Slater, A.F.A. Smit, E. Stupka, J. Szustakowki, D. Thierry-Mieg, J. Thierry-Mieg, L. Wagner, J. Wallis, R. Wheeler, A. Williams, Y.I. Wolf, K.H. Wolfe, S.P. Yang, R.F. Yeh, F. Collins, M.S. Guyer, J. Peterson, A. Felsenfeld, K.A. Wetterstrand, R.M. Myers, J. Schmutz, M. Dickson, J. Grimwood, D.R. Cox, M. V. Olson, R. Kaul, C. Raymond, N. Shimizu, K. Kawasaki, S. Minoshima, G.A. Evans, M. Athanasiou, R. Schultz, A. Patrinos, M.J. Morgan, Initial sequencing and analysis of the human genome, Nature 409 (2001) 860–921. 10.1038/35057062.

[3] C.R. Beck, P. Collier, C. Macfarlane, M. Malig, J.M. Kidd, E.E. Eichler, R.M. Badge, J. V. Moran, LINE-1 retrotransposition activity in human genomes, Cell 141 (2010) 1159–1170.

[4] B. Brouha, J. Schustak, R.M. Badge, S. Lutz-Prigge, A.H. Farley, J. V. Morant, H.H. Kazazian, Hot L1s account for the bulk of retrotransposition in the human population, Proc. Natl. Acad. Sci. U. S. A. 100 (2003) 5280–5285.

[5] S.J. Hoyt, J.M. Storer, G.A. Hartley, P.G.S. Grady, A. Gershman, L.G. de Lima, C. Limouse, R. Halabian, L. Wojenski, M. Rodriguez, N. Altemose, A. Rhie, L.J. Core, J.L. Gerton, W. Makalowski, D. Olson, J. Rosen, A.F.A. Smit, A.F. Straight, M.R. Vollger, T.J. Wheeler, M.C. Schatz, E.E. Eichler, A.M. Phillippy, W. Timp, K.H. Miga, R.J. O’Neill, From telomere to telomere: The transcriptional and epigenetic state of human repeat elements, Science. 376 (2022). 10.1126/science.abk3112.

[6] D.M. Sassaman, B.A. Dombroski, J. V. Moran, M.L. Kimberland, T.P. Naas, R.J. DeBerardinis, A. Gabriel, G.D. Swergold, H.H. Kazazian, Many human L1 elements are capable of retrotransposition, Nat. Genet. 16 (1997) 37–43.

[7] J.M.C. e. al Tubio, Extensive transduction of nonrepetitive DNA mediated by L1retrotransposition in cancer genomes, Science. 345 (2014) 1251343. 10.1126/science.1251343.Extensive.

[8] A.D. Ewing, A. Gacita, L.D. Wood, F. Ma, D. Xing, M.S. Kim, S.S. Manda, G. Abril, G. Pereira, A. Makohon-Moore, L.H.J. Looijenga, A.J.M. Gillis, R.H. Hruban, R.A. Anders, K.E. Romans, A. Pandey, C.A. Iacobuzio-Donahue, B. Vogelstein, K.W. Kinzler, H.H. Kazazian, S. Solyom, Widespread somatic L1 retrotransposition occurs early during gastrointestinal cancer evolution, Genome Res. 25 (2015) 1536–1545. 10.1101/gr.196238.115.

[9] B. Rodriguez-Martin, E.G. Alvarez, A. Baez-Ortega, J. Zamora, F. Supek, J. Demeulemeester, M. Santamarina, Y.S. Ju, J. Temes, D. Garcia-Souto, H. Detering, Y. Li, J. Rodriguez-Castro, A. Dueso-Barroso, A.L. Bruzos, S.C. Dentro, M.G. Blanco, G. Contino, D. Ardeljan, M. Tojo, N.D. Roberts, S. Zumalave, P.A.W. Edwards, J. Weischenfeldt, M. Puiggròs, Z. Chong, K. Chen, E.A. Lee, J.A. Wala, K. Raine, A. Butler, S.M. Waszak, F.C.P. Navarro, S.E. Schumacher, J. Monlong, F. Maura, N. Bolli, G. Bourque, M. Gerstein, P.J. Park, D.C. Wedge, R. Beroukhim, D. Torrents, J.O. Korbel, I.I. Martincorena, R.C. Fitzgerald, P. Van Loo, H.H. Kazazian, K.H. Burns, K.C. Akdemir, E.G. Alvarez, A. Baez-Ortega, R. Beroukhim, P.C. Boutros, D.D.L. Bowtell, B. Brors, K.H. Burns, P.J. Campbell, K. Chan, I. Cortés-Ciriano, A. Dueso-Barroso, A.J. Dunford, P.A.W. Edwards, X. Estivill, D. Etemadmoghadam, L. Feuerbach, J.L. Fink, M. Frenkel-Morgenstern, D.W. Garsed, M. Gerstein, D.A. Gordenin, D. Haan, J.E. Haber, J.M. Hess, B. Hutter, M. Imielinski, D.T.W. Jones, M.D. Kazanov, L.J. Klimczak, Y. Koh, J.O. Korbel, K. Kumar, E.A. Lee, J.J.K. Lee, Y. Li, A.G. Lynch, G. Macintyre, F. Markowetz, A. Martinez-Fundichely, M. Meyerson, S. Miyano, H. Nakagawa, F.C.P. Navarro, S. Ossowski, J. V. Pearson, J. V. Pearson, K. Rippe, N.D. Roberts, S.A. Roberts, B. Rodriguez-Martin, B. Rodriguez-Martin, S.E. Schumacher, M. Shackleton, N. Sidiropoulos, L. Sieverling, C. Stewart, J.M.C. Tubio, I. Villasante, N. Waddell, J.A. Wala, J. Weischenfeldt, L. Yang, X. Yao, S.S. Yoon, J. Zamora, C.Z. Zhang, P.J. Campbell, J.M.C. Tubio, S.E. Schumacher, R. Scully, B. Rodriguez-Martin, Y.S. Ju, M.D. Kazanov, L.J. Klimczak, Y. Koh, J.O. Korbel, K. Kumar, E.A. Lee, J.J.K. Lee, Y. Li, A.G. Lynch, G. Macintyre, F. Markowetz, I.I. Martincorena, A. Martinez-Fundichely, M. Meyerson, S. Miyano, H. Nakagawa, F.C.P. Navarro, S. Ossowski, J. V. Pearson, M. Puiggròs, K. Rippe, N.D. Roberts, S.A. Roberts, B. Rodriguez-Martin, S.E. Schumacher, R. Scully, M. Shackleton, N. Sidiropoulos, L. Sieverling, C. Stewart, J.M.C. Tubio, I. Villasante, N. Waddell, J.A. Wala, J. Weischenfeldt, L. Yang, X. Yao, S.S. Yoon, J. Zamora, C.Z. Zhang, P.J. Campbell, J.M.C. Tubio, Pan-cancer analysis of whole genomes identifies driver rearrangements promoted by LINE-1 retrotransposition, Nat. Genet. 52 (2020) 306–319. 10.1038/s41588-019-0562-0.

[10] A. Jou, J. Hess, Epidemiology and Molecular Biology of Head and Neck Cancer, Oncol. Res. Treat. 40 (2017) 328–332. 10.1159/000477127.

[11] M. Plath, J. Gass, M. Hlevnjak, Q. Li, B. Feng, X.P. Hostench, M. Bieg, L. Schroeder, D. Holzinger, M. Zapatka, K. Freier, W. Weichert, J. Hess, K. Zaoui, Unraveling most abundant mutational signatures in head and neck cancer, Int. J. Cancer 148 (2021) 115–127. 10.1002/IJC.33297.

[12] C.R. Leemans, B.J.M. Braakhuis, R.H. Brakenhoff, The molecular biology of head and neck cancer, Nat. Rev. Cancer 11 (2011) 9–22. 10.1038/nrc2982.

[13] H. Li, Aligning sequence reads, clone sequences and assembly contigs with BWA-MEM, ArXiv: Genomics (2013). 10.6084/M9.FIGSHARE.963153.V1.

[14] P. Danecek, J.K. Bonfield, J. Liddle, J. Marshall, V. Ohan, M.O. Pollard, A. Whitwham, T. Keane, S.A. McCarthy, R.M. Davies, Twelve years of SAMtools and BCFtools, Gigascience 10 (2021) 1–4. 10.1093/GIGASCIENCE/GIAB008.

[15] Broad Institute, Picard Tools, http://Broadinstitute.Github.Io/Picard/ (n.d.).

[16] W. Huang, L. Li, J.R. Myers, G.T. Marth, ART: a next-generation sequencing read simulator, Bioinformatics 28 (2012) 593–594. 10.1093/BIOINFORMATICS/BTR708.

[17] A.R. Quinlan, I.M. Hall, BEDTools: a flexible suite of utilities for comparing genomic features, Bioinformatics 26 (2010) 841–842. 10.1093/BIOINFORMATICS/BTQ033.

[18] M. V. Kuleshov, M.R. Jones, A.D. Rouillard, N.F. Fernandez, Q. Duan, Z. Wang, S. Koplev, S.L. Jenkins, K.M. Jagodnik, A. Lachmann, M.G. McDermott, C.D. Monteiro, G.W. Gundersen, A. Maayan, Enrichr: a comprehensive gene set enrichment analysis web server 2016 update, Nucleic Acids Res. 44 (2016) W90–W97. 10.1093/NAR/GKW377.

[19] D. Ardeljan, M.S. Taylor, D.T. Ting, K.H. Burns, The Human Long Interspersed Element-1 Retrotransposon: An Emerging Biomarker of Neoplasia, Clin. Chem. 63 (2017) 816–822. 10.1373/CLINCHEM.2016.257444.

[20] M.L. Filipenko, U.A. Boyarskikh, L.S. Leskov, K. V. Subbotina, E.A. Khrapov, A. V. Sokolov, I.S. Stilidi, N.E. Kushlinskii, The Level of LINE-1 mRNA Is Increased in Extracellular Circulating Plasma RNA in Patients with Colorectal Cancer, Bull. Exp. Biol. Med. 173 (2022) 261–264. 10.1007/S10517-022-05530-2/METRICS.

[21] S. Sato, M. Gillette, P.R. de Santiago, E. Kuhn, M. Burgess, K. Doucette, Y. Feng, C. Mendez-Dorantes, P.J. Ippoliti, S. Hobday, M.A. Mitchell, K. Doberstein, S.M. Gysler, M.S. Hirsch, L. Schwartz, M.J. Birrer, S.J. Skates, K.H. Burns, S.A. Carr, R. Drapkin, LINE-1 ORF1p as a candidate biomarker in high grade serous ovarian carcinoma, Sci. Rep. 13 (2023). 10.1038/s41598-023-28840-5.

[22] M.S. Taylor, C. Wu, P.C. Fridy, S.J. Zhang, Y. Senussi, J.C. Wolters, T. Cajuso, W.-C. Cheng, J.D. Heaps, B.D. Miller, K. Mori, L. Cohen, H. Jiang, K.R. Molloy, B.T. Chait, M.G. Goggins, I. Bhan, J.W. Franses, X. Yang, M.-E. Taplin, X. Wang, D.C. Christiani, B.E. Johnson, M. Meyerson, R. Uppaluri, A.M. Egloff, E.N. Denault, L.M. Spring, T.-L. Wang, I.-M. Shih, J.E. Fairman, E. Jung, K.S. Arora, O.H. Yilmaz, S. Cohen, T. Sharova, G. Chi, B.L. Norden, Y. Song, L.T. Nieman, L. Pappas, A.R. Parikh, M.R. Strickland, R.B. Corcoran, T. Mustelin, G. Eng, O.H. Yilmaz, U.A. Matulonis, S.J. Skates, B.R. Rueda, R. Drapkin, S.J. Klempner, V. Deshpande, D.T. Ting, M.P. Rout, J. LaCava, D.R. Walt, K.H. Burns, Ultrasensitive detection of circulating LINE-1 ORF1p as a specific multi-cancer biomarker, Cancer Discov. 13 (2023) OF1–OF16. 10.1158/2159-8290.CD-23-0313/729035/AM/ULTRASENSITIVE-DETECTION-OF-CIRCULATING-LINE-1.

[23] J.K. Baillie, M.W. Barnett, K.R. Upton, D.J. Gerhardt, T.A. Richmond, F. De Sapio, P.M. Brennan, P. Rizzu, S. Smith, M. Fell, R.T. Talbot, S. Gustincich, T.C. Freeman, J.S. Mattick, D.A. Hume, P. Heutink, P. Carninci, J.A. Jeddeloh, G.J. Faulkner, Somatic retrotransposition alters the genetic landscape of the human brain, Nature 479 (2011) 534–537. 10.1038/nature10531.

[24] C. Chu, R. Borges-Monroy, V. V. Viswanadham, S. Lee, H. Li, E.A. Lee, P.J. Park, Comprehensive identification of transposable element insertions using multiple sequencing technologies, Nat. Commun. 12 (2021). 10.1038/s41467-021-24041-8.

[25] E.J. Gardner, V.K. Lam, D.N. Harris, N.T. Chuang, E.C. Scott, W. Stephen Pittard, R.E. Mills, S.E. Devine, The mobile element locator tool (MELT): Population-scale mobile element discovery and biology, Genome Res. 27 (2017) 1916–1929. 10.1101/GR.218032.116/-/DC1.

[26] D.T. jwa. Thung, J. de Ligt, L.E.M. Vissers, M. Steehouwer, M. Kroon, P. de Vries, E.P. Slagboom, K. Ye, J.A. Veltman, J.Y. Hehir-Kwa, Mobster: accurate detection of mobile element insertions in next generation sequencing data, Genome Biol. 15 (2014) 488. 10.1186/S13059-014-0488-X/FIGURES/3.

[27] W. Zhou, S.B. Emery, D.A. Flasch, Y. Wang, K.Y. Kwan, J.M. Kidd, J. V. Moran, R.E. Mills, Identification and characterization of occult human-specific LINE-1 insertions using long-read sequencing technology, Nucleic Acids Res. 48 (2020) 1146–1163. 10.1093/NAR/GKZ1173.

[28] A.C. Katz-Summercorn, S. Jammula, A. Frangou, I. Peneva, M. O’Donovan, M. Tripathi, S. Malhotra, M. di Pietro, S. Abbas, G. Devonshire, W. Januszewicz, A. Blasko, K. Nowicki-Osuch, S. MacRae, A. Northrop, A.M. Redmond, D.C. Wedge, R.C. Fitzgerald, Multi-omic cross-sectional cohort study of pre-malignant Barrett’s esophagus reveals early structural variation and retrotransposon activity, Nat. Commun. 13 (2022). 10.1038/s41467-022-28237-4.

[29] A.W. Caliri, A. Caceres, S. Tommasi, A. Besaratinia, Hypomethylation of LINE-1 repeat elements and global loss of DNA hydroxymethylation in vapers and smokers, Epigenetics 15 (2020) 816–829. 10.1080/15592294.2020.1724401.

[30] H. Shigaki, Y. Baba, M. Watanabe, S. Iwagami, K. Miyake, T. Ishimoto, M. Iwatsuki, H. Baba, LINE-1 hypomethylation in noncancerous esophageal mucosae is associated with smoking history, Ann. Surg. Oncol. 19 (2012) 4238–4243. 10.1245/s10434-012-2488-y.

[31] P. Vineis, M. Alavanja, P. Buffler, E. Fontham, S. Franceschi, Y.T. Gao, P.C. Gupta, A. Hackshaw, E. Matos, J. Samet, F. Sitas, J. Smith, L. Stayner, K. Straif, M.J. Thun, H.E. Wichmann, A.H. Wu, D. Zaridze, R. Peto, R. Doll, Tobacco and cancer: recent epidemiological evidence, J. Natl. Cancer Inst. 96 (2004) 99–106. 10.1093/JNCI/DJH014.

[32] K. Hur, P. Cejas, J. Feliu, J. Moreno-Rubio, E. Burgos, C.R. Boland, A. Goel, Hypomethylation of long interspersed nuclear element-1 (LINE-1) leads to activation of proto-oncogenes in human colorectal cancer metastasis, Gut 63 (2014) 635–646. 10.1136/GUTJNL-2012-304219.

[33] M. Casarotto, V. Lupato, G. Giurato, R. Guerrieri, S. Sulfaro, A. Salvati, E. D’Angelo, C. Furlan, A. Menegaldo, L. Baboci, B. Montico, I. Turturici, R. Dolcetti, S. Romeo, V. Baggio, S. Corrado, G. Businello, M. Guido, A. Weisz, V. Giacomarra, G. Franchin, A. Steffan, L. Sigalotti, E. Vaccher, P. Boscolo-Rizzo, P. Jerry, G. Fanetti, E. Fratta, LINE-1 hypomethylation is associated with poor outcomes in locoregionally advanced oropharyngeal cancer, Clin. Epigenetics 14 (2022). 10.1186/s13148-022-01386-5.

[34] C. Furlan, J. Polesel, L. Barzan, G. Franchin, S. Sulfaro, S. Romeo, F. Colizzi, A. Rizzo, V. Baggio, V. Giacomarra, A.P. Dei Tos, P. Boscolo-Rizzo, E. Vaccher, R. Dolcetti, L. Sigalotti, E. Fratta, Prognostic significance of LINE-1 hypomethylation in oropharyngeal squamous cell carcinoma, Clin. Epigenetics 9 (2017). 10.1186/S13148-017-0357-Z/FIGURES/4.

[35] K. Misawa, S. Yamada, M. Mima, T. Nakagawa, T. Kurokawa, A. Imai, D. Mochizuki, D. Shinmura, T. Yamada, J. Kita, R. Ishikawa, Y. Yamaguchi, Y. Misawa, T. Kanazawa, H. Kawasaki, H. Mineta, Long interspersed nuclear element 1 hypomethylation has novel prognostic value and potential utility in liquid biopsy for oral cavity cancer, Biomark. Res. 8 (2020) 1–10. 10.1186/S40364-020-00235-Y/FIGURES/6.

[36] A. Benard, C.J.H. Van De Velde, L. Lessard, H. Putter, L. Takeshima, P.J.K. Kuppen, D.S.B. Hoon, Epigenetic status of LINE-1 predicts clinical outcome in early-stage rectal cancer, Br. J. Cancer 109 (2013) 3073–3083. 10.1038/bjc.2013.654.

[37] E.J. Kim, W.C. Chung, D.B. Kim, Y.J. Kim, J.M. Lee, J.H. Jung, Y.K. Lee, Long interspersed nuclear element (LINE)-1 methylation level as a molecular marker of early gastric cancer, Dig. Liver Dis. 48 (2016) 1093–1097. 10.1016/j.dld.2016.06.002.

[38] E. Sunami, M. de Maat, A. Vu, R.R. Turner, D.S.B. Hoon, LINE-1 Hypomethylation During Primary Colon Cancer Progression, PLoS One 6 (2011) e18884. 10.1371/JOURNAL.PONE.0018884.

[39] J.P. Foy, C.R. Pickering, V.A. Papadimitrakopoulou, J. Jelinek, S.H. Lin, W.N. William, M.J. Frederick, J. Wang, W. Lang, L. Feng, L. Zhang, E.S. Kim, Y.H. Fan, W.K. Hong, A.K. El-Naggar, J.J. Lee, J.N. Myers, J.P. Issa, S.M. Lippman, L. Mao, P. Saintigny, New DNA methylation markers and global DNA hypomethylation are associated with oral cancer development, Cancer Prev. Res. 8 (2015) 1027–1035. 10.1158/1940-6207.CAPR-14-0179.

[40] W. McKerrow, X. Wang, C. Mendez-Dorantes, P. Mita, S. Cao, M. Grivainis, L. Ding, J. LaCava, K.H. Burns, J.D. Boeke, D. Fenyö, LINE-1 expression in cancer correlates with p53 mutation, copy number alteration, and S phase checkpoint, Proc. Natl. Acad. Sci. U. S. A. 119 (2022). 10.1073/PNAS.2115999119/-/DCSUPPLEMENTAL.

[41] B. Tiwari, A.E. Jones, C.J. Caillet, S. Das, S.K. Royer, J.M. Abrams, P53 directly represses human LINE1 transposons, Genes Dev. 34 (2020) 1439–1451. 10.1101/GAD.343186.120/-/DC1.

[42] P. Mita, X. Sun, D. Fenyö, D.J. Kahler, D. Li, N. Agmon, A. Wudzinska, S. Keegan, J.S. Bader, C. Yun, J.D. Boeke, BRCA1 and S phase DNA repair pathways restrict LINE-1 retrotransposition in human cells, Nat. Struct. Mol. Biol. 27 (2020) 179–191. 10.1038/S41594-020-0374-Z.

[43] N. Rodić, R. Sharma, R. Sharma, J. Zampella, L. Dai, M.S. Taylor, R.H. Hruban, C.A. Iacobuzio-Donahue, A. Maitra, M.S. Torbenson, M. Goggins, I.M. Shih, A.S. Duffield, E.A. Montgomery, E. Gabrielson, G.J. Netto, T.L. Lotan, A.M. De Marzo, W. Westra, Z.A. Binder, B.A. Orr, G.L. Gallia, C.G. Eberhart, J.D. Boeke, C.R. Harris, K.H. Burns, Long interspersed element-1 protein expression is a hallmark of many human cancers, Am. J. Pathol. 184 (2014) 1280–1286. 10.1016/j.ajpath.2014.01.007.

[44] D. Mangoni, A. Simi, P. Lau, A. Armaos, F. Ansaloni, A. Codino, D. Damiani, L. Floreani, V. Di Carlo, D. Vozzi, F. Persichetti, C. Santoro, L. Pandolfini, G.G. Tartaglia, R. Sanges, S. Gustincich, LINE-1 regulates cortical development by acting as long non-coding RNAs, Nat. Commun. 14 (2023). 10.1038/s41467-023-40743-7.

[45] M. Martínez-Fernández, A. Feber, M. Dueñas, C. Segovia, C. Rubio, M. Fernandez, F. Villacampa, J. Duarte, F.F. López-Calderón, M.J. Gómez-Rodriguez, D. Castellano, J.L. Rodriguez-Peralto, F. De La Rosa, S. Beck, J.M. Paramio, Analysis of the polycomb-related lncRNAs HOTAIR and ANRIL in bladder cancer, Clin. Epigenetics 7 (2015). 10.1186/s13148-015-0141-x.

[46] C.A. Ishak, A.E. Marshall, D.T. Passos, C.R. White, J. Seung, M.J. Cecchini, S. Ferwati, W.A. Macdonald, J. Christopher, I.D. Welch, S.M. Rubin, M.R.W. Mann, F.A. Dick, An RB-EZH2 Complex Mediates Silencing of Repetitive DNA Sequences, 64 (2017) 1074–1087. 10.1016/j.molcel.2016.10.021.An.

[47] P. V. Angadi, J.K. Savitha, S.S. Rao, R.Y. Sivaranjini, Oral field cancerization: Current evidence and future perspectives, Oral Maxillofac. Surg. 16 (2012) 171–180. 10.1007/S10006-012-0317-X/TABLES/1.

[48] I. Martincorena, A. Roshan, M. Gerstung, P. Ellis, P. Van Loo, S. McLaren, D.C. Wedge, A. Fullam, L.B. Alexandrov, J.M. Tubio, L. Stebbings, A. Menzies, S. Widaa, M.R. Stratton, P.H. Jones, P.J. Campbell, High burden and pervasive positive selection of somatic mutations in normal human skin, Science. 348 (2015) 880–886. 10.1126/SCIENCE.AAA6806/SUPPL_FILE/AAA6806-MARTINCORENA-SM.PDF.

[49] K. Yoshida, K.H.C. Gowers, H. Lee-Six, D.P. Chandrasekharan, T. Coorens, E.F. Maughan, K. Beal, A. Menzies, F.R. Millar, E. Anderson, S.E. Clarke, A. Pennycuick, R.M. Thakrar, C.R. Butler, N. Kakiuchi, T. Hirano, R.E. Hynds, M.R. Stratton, I. Martincorena, S.M. Janes, P.J. Campbell, Tobacco smoking and somatic mutations in human bronchial epithelium, Nat. 2020 5787794 578 (2020) 266–272. 10.1038/s41586-020-1961-1.

[50] T.G. Paulson, P.C. Galipeau, K.M. Oman, C.A. Sanchez, M.K. Kuhner, L.P. Smith, K. Hadi, M. Shah, K. Arora, J. Shelton, M. Johnson, A. Corvelo, C.C. Maley, X. Yao, R. Sanghvi, E. Venturini, A.K. Emde, B. Hubert, M. Imielinski, N. Robine, B.J. Reid, X. Li, Somatic whole genome dynamics of precancer in Barrett’s esophagus reveals features associated with disease progression, Nat. Commun. 13 (2022). 10.1038/s41467-022-29767-7.

[51] I. Martincorena, J.C. Fowler, A. Wabik, A.R.J. Lawson, F. Abascal, M.W.J. Hall, Cagan, K. Murai, K. Mahbubani, M.R. Stratton, R.C. Fitzgerald, P.A. Handford, P.J. Campbell, K. Saeb-Parsy, P.H. Jones, Somatic mutant clones colonize the human esophagus with age, Science. 362 (2018) 911–917. 10.1126/science.aau3879.

[52] A.R.J. Lawson, F. Abascal, T.H.H. Coorens, Y. Hooks, L. O’Neill, C. Latimer, K. Raine, M.A. Sanders, A.Y. Warren, K.T.A. Mahbubani, B. Bareham, T.M. Butler, L.M.R. Harvey, A. Cagan, A. Menzies, L. Moore, A.J. Colquhoun, W. Turner, B. Thomas, V. Gnanapragasam, N. Williams, D.M. Rassl, H. Vöhringer, S. Zumalave, J. Nangalia, J.M.C. Tubío, M. Gerstung, K. Saeb-Parsy, M.R. Stratton, P.J. Campbell, T.J. Mitchell, I. Martincorena, Extensive heterogeneity in somatic mutation and selection in the human bladder, Science. 370 (2020) 75–82. 10.1126/science.aba8347.

[53] C.H. Nam, J. Youk, J.Y. Kim, J. Lim, J.W. Park, S.A. Oh, H.J. Lee, J.W. Park, H. Won, Y. Lee, S.Y. Jeong, D.S. Lee, J.W. Oh, J. Han, J. Lee, H.W. Kwon, M.J. Kim, Y.S. Ju, Widespread somatic L1 retrotransposition in normal colorectal epithelium, Nature 617 (2023) 540–547. 10.1038/s41586-023-06046-z.

[54] M. Shademan, K. Zare, M. Zahedi, H. Mosannen Mozaffari, H. Bagheri Hosseini, K. Ghaffarzadegan, L. Goshayeshi, H. Dehghani, Promoter methylation, transcription, and retrotransposition of LINE-1 in colorectal adenomas and adenocarcinomas, Cancer Cell Int. 20 (2020) 1–16. 10.1186/S12935-020-01511-5.

