## Additional file 1 for "RetroTest unravels LINE-1 retrotransposition in Head and Neck Squamous Cell Carcinoma"

### SUPPLEMENTARY METHODS

#### RetroTest library and target sequencing

Sequencing libraries enriched in L1 3'-transductions were prepared starting from 100ng of DNA sheared using a Covaris M220 Focused-Ultrasonicator (Covaris Inc.) in fragments of ~300bp for FFPE and ~500bp for frozen tumors. After sonication, fragment size and DNA concentration were assessed with High Sentitivity DNA Assay (Agilent Technologies Inc.). Adaptor-ligated libraries of HNSCC samples were prepared using SureSelect XT2 Library Prep Kit. Briefly, the samples were indexed, amplified, and pooled before hybridization and capture with RNA targeted baits. Captured indexed pools were amplified to obtain final enriched libraries. Libraries of lung and bladder cancer samples were prepared with SureSelect XT HS Target Enrichment using the automated Magnis NGS Prep system (Agilent Technologies Inc.), where fragmented samples after Covaris sonication were loaded to obtain final enriched libraries. Quality controls of the library preparation were performed by using D1000 ScreenTape Assay and High Sensitivity D1000 Assay (Agilent Technologies Inc.). The multiplexed samples were sequenced on Illumina MiSeq and NextSeq platforms using pair-end reads with MiSeq Reagent Kit v2 and NextSeq 500/550 Mid Output Kit v2.5 (300 Cycles) (Illumina Inc.).

#### RetroTest method in detail

The input of RetroTest is processed BAM files with 150bp reads, derived from Illumina paired-end sequencing. RetroTest can be run in either tumor-only or tumor-normal matched modes. The identification of insertion supporting clusters is performed as follows:

(i) Search for discordant and clipped read events in the BAM file/s. This step can be performed in a. Single sample mode or b. Tumor-normal matched mode if a germline control is provided. Then, left- and right-clipped read events are realigned to search for supplementary alignments.

(ii) Discordant and clipping read events are organized into genomic bins and then grouped into clusters. Then discordant read pairs are grouped based on mate position while clipping read events are grouped based on the supplementary alignment position. The genomic bins to search for insertions (will correspond to transduced areas) are based on the coordinates of the L1 downstream-transduced regions.

(iii) For cluster filtering, all clusters without the minimum number of reads per cluster, clusters in unspecific regions, and clusters composed of duplicated reads are discarded. Clusters can also be filtered out based on supplied genomic coordinates and average alignment mapping quality. Additionally, for discordant clusters, those with mates not over the target reference, and those whose mates align over any source element downstream region are discarded. For clipping clusters, those with a supplementary alignment outside the target reference and those whose supplementary alignments map over any source element downstream region are also filtered out.

(iv) Filtered discordant and clipping clusters are grouped into metaclusters, whose precise coordinates are determined by metacluster breakpoints. Their precise coordinates are determined by the metacluster breakpoints.

(v) Finally, each metacluster transduction type is determined using the discordant reads around the insertion point, based on the mapping position of the anchor's mate.

The pipeline can be found in the following site:

https://gitlab.com/mobilegenomesgroup/RETROTEST

### RetroTest benchmarking

To simulate orphan transductions, 5Kbp of downstream reference genome (hg19) sequence was retrieved for each of the 124 source elements included in MEIGA-MEIsimulator database[1], after considering L1 orientation. For each source element, 20 transduction sequences were generated by random 3' trimming, representing alternative transcription endings. To simulate the insertion events, reference genome sequence was divided into 10kb bins, and insertion points were randomly selected among the bins contained by nuclear chromosomes, with the only condition of avoiding GAP regions. DNA sequences from reference genome and orphan transductions were sequentially merged using custom python commands, while adding MEI characteristic features, including target site duplication, polyA tail and 5' truncation.

In silico paired-end reads from this modified genome version were generated with ART v2.5.8 (150bp length, insert size 350bp ± 10%) and subsequently aligned with BWA-mem v0.7.17[2,3] against the reference genome, and further processed with samtools v1.3.1[3,4].

### Whole Genome Sequencing and determination of mutation profile

Sequencing reads from tumor tissue and normal adjacent tissue (NAT) were mapped to the hg19 reference genome by Burrows-Wheeler Aligner BWA-mem[2,3] v0.7.17. Samtools[4,5] v 1.9 was used to sort the aligned reads and to index the obtained bam file, applying Picard Bammarkduplicates2[6] to mark duplicated reads. After that, Mutect2[7], from the Genome Analysis Tool Kit (GATK)[8] v4.1.1.0, was used to perform SNVs and INDELs calling. Variants were filtered with FilterMutectCalls (GATK) (considering ASCAT[9] normal contamination estimation), following standard thresholds, and annotated using the Ensembl Variant Effect Predictor (VEP)[10] v100.2. We selected those probably pathogenic variants, following SIFT[11,12], PolyPhen[13], and VEP Impact annotations. Then, we ensured our variants were somatic by filtering those with an allele frequency equal to or higher than 0.01 in the 1KGP, ESP, or genomAD populations, assessing they are not common in the population.

To analyze the mutational profile of normal adjacent tissue (NAT), we also sequenced blood for 4 patients. We used again Mutect2 v4.1.7.0 to perform joint SNVs and INDELs calling both for tumor and NAT, following best practices[8,14]. In this case, GATK FilterMutectCalls now considered cross-sample contamination estimates performed by GATK CalculateContamination. We considered as common between NAT and tumor those variants presenting more than one supporting read in both samples, while those exclusive from NAT presented more than one supporting read in this sample and less than 1 in the tumor (normally, 0).

### Gene pathway databases uses for enrichment analysis

Human MSigDB collections[15,16] (MSigDB_Hallmark_2020, MSigDB_Oncogenic_Signatures, and MSigDB_Computational) BioPlanet_2019[17], KEGG_2019_Human[18,19], WikiPathways_2019_Human[20,21], GO_Molecular_Function_2018, and GO_Biological_Process_2018[22,23]. We used the following transcription factor binding motifs databases: TRANSFAC_and_JASPAR_PWMs[24] and ChEA_2016[25,26].

### REFERENCES


1. Ebert, P. *et al.* Haplotype-resolved diverse human genomes and integrated analysis of structural variation. *Science* **372**, (2021).

2. Li, H. Aligning sequence reads, clone sequences and assembly contigs with BWA-MEM. *arXiv: Genomics* (2013). doi:10.6084/M9.FIGSHARE.963153.V1

3. Li, H. & Durbin, R. Fast and accurate short read alignment with Burrows-Wheeler transform. *Bioinformatics* **25**, 1754–1760 (2009).

4. Danecek, P. *et al.* Twelve years of SAMtools and BCFtools. *Gigascience* **10**, 1–4 (2021).

5. Li, H. *et al.* The Sequence Alignment/Map format and SAMtools. *Bioinformatics* **25**, 2078–2079 (2009).

6. Broad Institute. Picard Tools. *http://broadinstitute.github.io/picard/*

7. Benjamin, D. *et al.* Calling Somatic SNVs and Indels with Mutect2. *bioRxiv* 861054 (2019). doi:10.1101/861054

8. Van der Auwera, G. A. & O'Connor, B. D. *Genomics in the Cloud: Using Docker, GATK, and WDL in Terra (1st Edition)*. (O'Reilly Media, 2020).

9. Van Loo, P. *et al.* Allele-specific copy number analysis of tumors. *Proc. Natl. Acad. Sci. U. S. A.* **107**, 16910–16915 (2010).

10. McLaren, W. *et al.* The Ensembl Variant Effect Predictor. *Genome Biol.* **17**, 1–14 (2016).

11. Kumar, P., Henikoff, S. & Ng, P. C. Predicting the effects of coding non-synonymous variants on protein function using the SIFT algorithm. *Nat. Protoc. 2009 47* **4**, 1073–1081 (2009).

12. Sim, N. L. *et al.* SIFT web server: predicting effects of amino acid substitutions on proteins. *Nucleic Acids Res.* **40**, W452–W457 (2012).

13. Adzhubei, I. A. *et al.* A method and server for predicting damaging missense mutations. *Nat. Methods* **7**, 248–249 (2010).

14. GATK Broad Institute & Caetano-Anolles, D. Somatic short variant discovery (SNVs + Indels). *https://gatk.broadinstitute.org/hc/en-us/articles/360035894731-Somatic-short-variant-discovery-SNVs-Indels-*

15. Liberzon, A. *et al.* The Molecular Signatures Database (MSigDB) hallmark gene set collection. *Cell Syst.* **1**, 417 (2015).

16. Subramanian, A. *et al.* Gene set enrichment analysis: A knowledge-based approach for interpreting genome-wide expression profiles. *Proc. Natl. Acad. Sci. U. S. A.* **102**, 15545–15550 (2005).

17. Huang, R. *et al.* The NCATS BioPlanet – An integrated platform for exploring the universe of cellular signaling pathways for toxicology, systems biology, and chemical genomics. *Front. Pharmacol.* **10**, 437284 (2019).

18. Kanehisa, M. Toward understanding the origin and evolution of cellular organisms. *Protein Sci.* **28**, 1947–1951 (2019).

19. Kanehisa, M., Furumichi, M., Sato, Y., Kawashima, M. & Ishiguro-Watanabe, M. KEGG for taxonomy-based analysis of pathways and genomes. *Nucleic Acids Res.* **51**, D587–D592 (2023).

20. Martens, M. *et al.* WikiPathways: connecting communities. *Nucleic Acids Res.* **49**, D613–D621 (2021).

21. Pico, A. R. *et al.* WikiPathways: Pathway Editing for the People. *PLOS Biol.* **6**, e184 (2008).

22. Ashburner, M. *et al.* Gene Ontology: tool for the unification of biology. *Nat. Genet. 2000 251* **25**, 25–29 (2000).

23. Consortium, T. G. O. *et al.* The Gene Ontology knowledgebase in 2023. *Genetics* **224**, (2023).

24. Fornes, O. *et al.* JASPAR 2020: update of the open-access database of transcription factor binding profiles. *Nucleic Acids Res.* **48**, D87–D92 (2020).

25. Kuleshov, M. V. *et al.* Enrichr: a comprehensive gene set enrichment analysis web

server 2016 update. *Nucleic Acids Res.* **44**, W90–W97 (2016).

26. Xie, Z. *et al.* Gene Set Knowledge Discovery with Enrichr. *Curr. Protoc.* **1**, e90 (2021).
