## Additional file 3 for "RetroTest unravels LINE-1 retrotransposition in Head and Neck Squamous Cell Carcinoma"

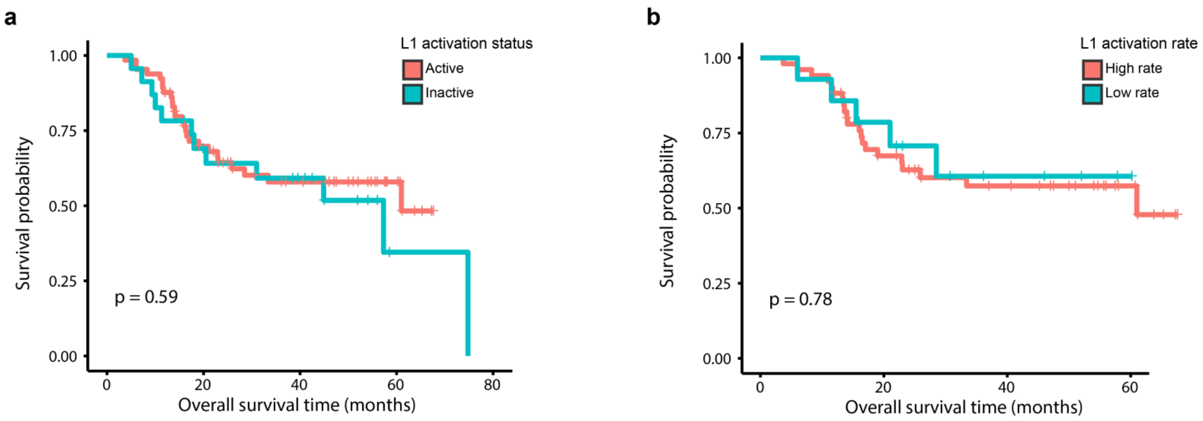


**Figure S1.** Kaplan-Meier curves for overall survival in HNSCC cohort with respect to (A) L1 activation status (active vs inactive) and (B) L1 activation rate (with respect to the median, being high above vs low as below the median). Log-rank test was used to calculate the p-value.
